# Detection of Carbapenem-Resistant Bacteria in Skilled Nursing Facility Wastewater

**DOI:** 10.1101/2025.07.17.25331724

**Authors:** Susanna Lenz, Subhasish Biswas, Ivy Terry, Lisa Tran Basha, Amanda K. Lyons, Tapati Mazumdar, Florence Whitehill, Alison Laufer Halpin, Lauren Franco, Maria Karlsson, Margaret Williams, Angela Coulliette-Salmond

## Abstract

Wastewater surveillance at healthcare facilities for carbapenem-resistant (CR) bacteria harboring carbapenemase genes could result in earlier detection of circulating or emerging antimicrobial resistance (AR) threats. However, knowledge gaps exist regarding the CR bacterial community and carbapenemase genes present in facility wastewater and plumbing, which need to be addressed. Wastewater effluent samples from three skilled nursing facilities (SNFs) (Georgia, USA) were collected weekly over six months each. Selective chromogenic media were examined for their performance in screening the samples for targeted CR bacteria. Culture-dependent and culture-independent methods (i.e., PCR) were applied to enriched wastewater samples (n=73) to screen isolates and pooled samples for the most common carbapenemase genes circulating in the United States (*bla*_KPC_, *bla*_NDM_, *bla*_VIM_, *bla*_OXA-48-like_, and *bla*_IMP_), and method sensitivity was compared. Whole genome sequencing (WGS) was used to identify all AR genes present in isolates. Bacteria (n=209 isolates) were identified using MALDI-TOF and included >20 different genera. Clinically relevant (*Acinetobacter baumannii* complex, *Enterobacter cloacae* complex, *Enterococcus faecium, Stenotrophomonas* spp.) and environmental/opportunistic (*Comamonas* spp., *Pandoraea* spp*., Pseudomonas putida* group) CR bacteria were identified in each of the three SNFs. In Facility A and C, only *bla*_KPC_ was detected in 9 (26.5%) of 34 and 14 (11.4%) of 123 recovered isolates, respectively. There were no PCR-positive results in Facility B. WGS confirmed the presence or absence of all genes screened for by PCR (100% concordance). These data provide a proof of concept and insights into CR bacteria present in healthcare facility wastewater using culture, PCR, and sequencing methods.

## Introduction

Carbapenem-resistant Enterobacteriaceae (CRE) are responsible for an estimated 13,100 infections in hospitalized patients and an estimated $130 million in attributable healthcare costs based on 2017 data [1]. CRE are particularly concerning in health facilities that care for high-acuity patients with long lengths of stay (>30 days), such as ventilator-capable skilled nursing facilities (vSNFs) [1, 2]. vSNFs are considered ‘influential facilities’ due to their substantial impact on regional transmission of antimicrobial resistance (AR); models demonstrate that earlier detection in vSNFs with implementation of targeted infection control practices would reduce regional spread overall [3–6]. Currently, prevention-driven admission screening and point prevalence surveys (PPSs), both periodic and following identification of any vSNF resident with a novel or targeted multidrug-resistant organism, are two detection strategies to help prevent the spread of CRE and other AR/carbapenem-resistant (CR) bacteria [7]. However, limited staffing, resources, and other capacity limitations can impede these approaches.

Given these challenges, wastewater surveillance (WWS) offers a more feasible approach for the detection of antimicrobial-resistant bacteria from healthcare facilities, as shown by previous studies and reports [8–13]. WWS at vSNFs could have an exponential impact, protecting not only the individual facility’s population but also patients at facilities in each vSNF’s transfer network. However, WWS work in vSNFs, particularly for CR bacteria, is limited, and robust methodological comparisons and antimicrobial-resistant bacteria baselining efforts are lacking. One pilot study demonstrated successful detection of CRE at a SNF, but did not compare the wastewater data with clinical surveillance [14], a critical component for leveraging WWS as a tool for infection prevention and control.

In the United States, the Centers for Disease Control and Prevention (CDC) AR Laboratory Network (AR Lab Network) is designed to detect and respond to healthcare-associated infection/AR threats through clinical testing, including PPSs; however, healthcare facility WWS data are lacking. Furthermore, inconsistencies and weak correlations with community-level WWS for AR and population clinical surveillance metrics have been reported [13, 15, 16]. To address these gaps and further healthcare WWS implementation efforts in the United States, this longitudinal WWS study aimed to characterize the CR bacteria baseline detected at healthcare facilities and to compare and harmonize existing environmental and clinical methods for use in wastewater. Of importance was to align the traditional environmental method (i.e., IDEXX Colilert-18) with clinical methods used by AR Lab Network public health laboratories [17] to facilitate correlations between WWS and patient testing results. The project was conducted at three SNFs, which were already participating in a separate, ongoing surveillance effort [14]. Although these SNFs do not serve as influential facilities or offer ventilator care, they were considered a suitable proxy for the study’s objectives. The AR/bacterial targets of this study were carbapenem-resistant *Acinetobacter* spp., *Pseudomonas* spp., and CRE, and the most common types of carbapenemases: *Klebsiella pneumoniae* carbapenemases (KPC), oxacillinase (OXA)-48-like β-lactamases, and the metallo-β-lactamases New Delhi metallo-β-lactamases (NDM), active-on-imipenem family of carbapenemases (IMP), and Verona integron-encoded metallo-β-lactamases (VIM).

## Methods

This effort met the US Centers for Disease Control and Prevention (CDC) safety policies and recommendations outlined in the CDC/NIH *Biosafety in Microbiological and Biomedical Laboratories 17* [18]. This activity was reviewed by the CDC and was conducted consistent with applicable federal law and CDC policy*§*.

### Facilities and Wastewater Sampling

Three SNFs in the Atlanta-metro area (Georgia, USA), herein referred to as Facility A, B, and C, participated in this wastewater surveillance project during 2022 for six months, or until 25 weekly samples were collected. Facility A and B provided skilled nursing, inpatient rehabilitation, wound/ostomy, and indwelling device types of care; Facility C provided similar care as well as tracheostomy care. The median census for each facility was determined by facility self-reporting during wastewater sampling periods (Facility A: February to July 2022; Facility B: March to September 2022; Facility C: June to December 2022).

Twenty-four-hour composite samples were collected weekly from a manhole located on each property, closest to the SNF building, using an Avalanche Portable Refrigerated Autosampler (Teledyne ISCO, Lincoln, NE, USA). For further details regarding field sampling methods and considerations, refer to Santiago et al. 2025 [14] and Coulliette-Salmond et al. 2025 [19].

### *Wastewater Enrichment and* E. coli *Detection Using Colilert-18 Assay*

Collected wastewater samples were screened for *Escherichia coli (E. coli)* using the Colilert-18 assay (IDEXX Laboratories, Inc., Westbrook, ME, USA), and the concentration of viable *E. coli* was estimated using the most probable number (MPN) method, according to manufacturer instructions. The foil-side of the Colilert-18 trays were cleaned with 70% ethanol for aseptic technique, and a pooled sample (1 mL total) was pulled from five or 20% of randomly selected *E. coli*-positive large wells per tray (which ever number was larger) using a three mL needle syringe to pull at least 100 µL per well [20–23]. A total of 73 pooled samples were collected (two samples from Facility A were unavailable for downstream processing) and stored at −80°C in 25% glycerol for downstream analysis of AR genes for both the ‘enrichment-to-isolate’ and ‘enrichment-to-PCR’ workflows (Figure 1).

**Figure 1:**
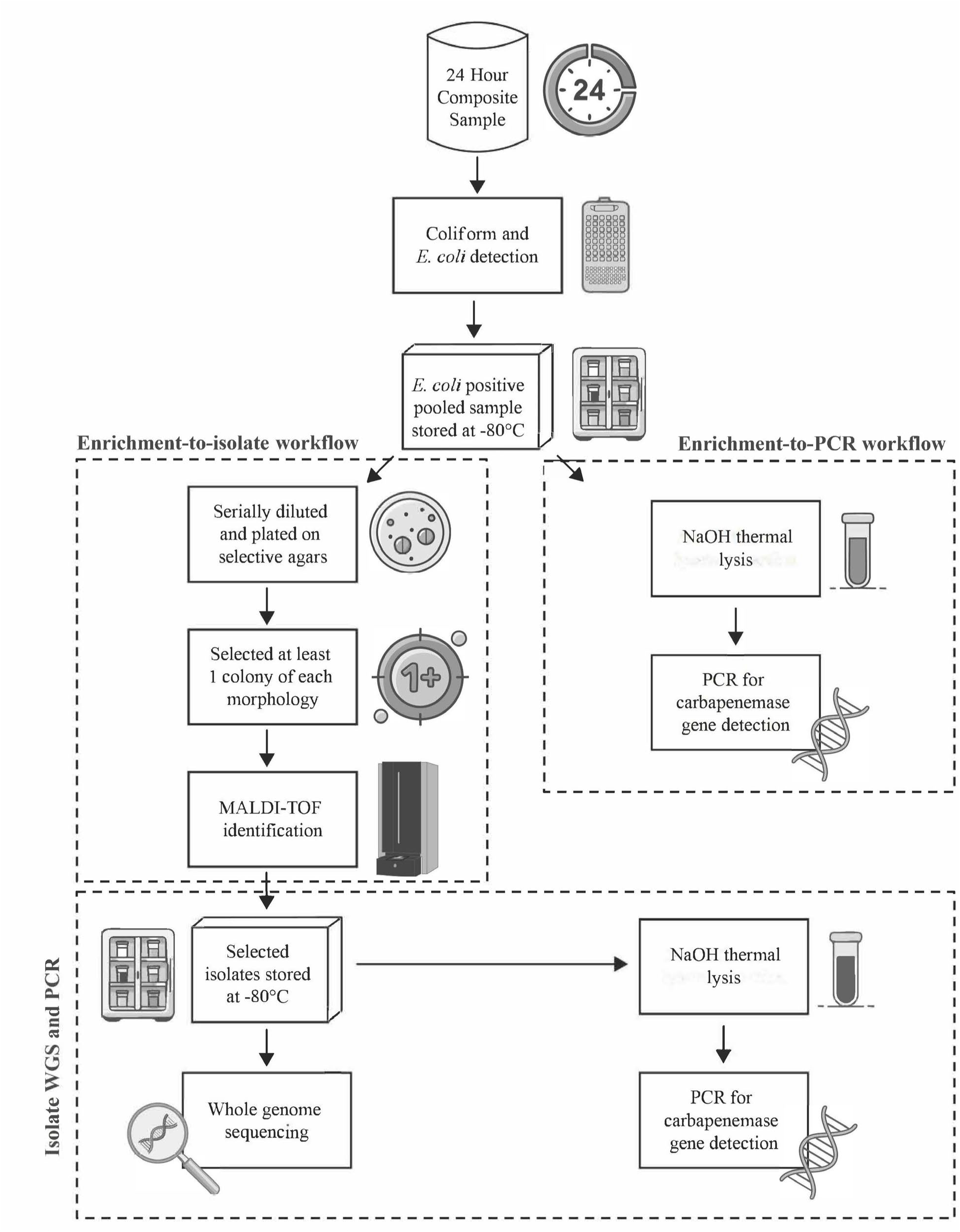
Experimental workflow for enriching, isolating, and detecting carbapenem-resistant bacteria and carbapenemase genes from wastewater samples at three skilled nursing facilities (Georgia, USA; 2022).

### Culture-based Detection of Carbapenem-Resistant Bacteria

*Enrichment-to-isolate workflow*: *E. coli*-positive Colilert-18 pooled samples were screened for CRE using selective chromogenic media, CHROMagar mSuperCARBA (mSC) (DRG International, Springfield, NJ USA). Three additional agars were used to further evaluate the presence of other carbapenem-resistant bacteria, including those displaying intrinsic resistance: CHROMagar Acinetobacter + MDR Supplement (AC) (DRG International, Springfield, NJ, USA), CHROMagar Pseudomonas (PC) (DRG International, Springfield, NJ, USA), and MacConkey agar with 10 µg ertapenem and meropenem disks (MAC+erta, MAC+mero). For MAC+erta and MAC+mero, one MacConkey agar plate was divided in half, and one disk of each antibiotic was placed on each half. Colonies inside the zone of inhibition were chosen for identification.

Pooled samples were thawed, serially diluted, and 100 µL of each dilution was spread-plated on mSC, AC, PC, and MAC+erta+mero and incubated overnight at 35°C or 30°C, according to product inserts. For each of the 73 samples evaluated, at least one colony of each morphology per agar (i.e., convenience subset of all colonies) was selected for identification using Matrix-Assisted Laser Desorption/Ionization Time-of-Flight (MALDI-TOF; MALDI Biotyper, Bruker Daltonics, Billerica, MA, USA) with the Bruker and CDC MicrobeNet databases (https://microbenet.cdc.gov). Following identification and documentation of confidence scores (species level: 2.000-3.000; genera level: 1.700-1.999), freezer stocks of isolates of interest were stored at −80°C (cryobank tubes Mast Group Ltd., Merseyside, UK).

*Classification*: Detected species were classified into two groups: clinically relevant or environmental/opportunistic pathogens. Groupings were primarily based on clinical significance as described in the Manual of Clinical Microbiology [24]. Five additional species were deemed clinically relevant based on input from clinical microbiologists and observations from the CDC’s healthcare-associated pathogen reference and surveillance testing (personal communication – Maria Karlsson, Lauren Franco): *Aeromonas hydrophila*, *Burkholderia cepacia*, *Citrobacter freundii*, *Stenotrophomonas maltophilia*, and *Serratia marcescens*. A threshold percentage to define ‘frequently detected’ bacteria was set a priori at 25%, which translates to detection in six or more wastewater composite samples from each facility.

*Enrichment-to-PCR workflow*: *E. coli*-positive Colilert-18 wastewater pooled samples were aliquoted (termed “pooled lysate”), and a 1 mL sample was extracted using CDC’s thermal NaOH bacterial lysate method (https://www.cdc.gov/gram-negative-bacteria/php/laboratories/). Lysates were frozen (−20°C) until multiplex real-time PCR was performed for the *bla*_KPC_, *bla*_NDM_ (https://www.cdc.gov/gram-negative-bacteria/php/laboratories/), *bla*_VIM_, *bla*_OXA-48-like_ [25], and *bla*_IMP_ [26] carbapenemase genes using assays previously described. Select samples were screened for *bla*_OXA-23_ [27] due to a facility having a resident with confirmed *bla*_OXA-23_-positive *A. baumannii* infection.

### Molecular-based Detection of Carbapenemase Genes

*PCR workflow:* All bacterial isolates were grown on TSA + 5% sheep blood agar, and then a 1 µL-loopful of bacterial growth was extracted using the Thermal NaOH Bacterial Lysate method. Extracts were frozen (−20°C) until multiplex real-time PCR was performed for the *bla*_KPC_, *bla*_NDM_, *bla*_VIM_, *bla*_OXA-48-like_, and *bla*_IMP_ carbapenemase genes using CDC assays previously described.

*Whole genome sequencing workflow:* Isolates (n=182) were chosen for whole genome sequencing (WGS) based on several criteria: all PCR-positive isolates, and one organism per unique species from each wastewater composite sample. If the same species was isolated from multiple agar types within a single composite sample, isolates cultured from mSC were prioritized for sequencing. DNA was extracted from isolates using the Promega RSC Cultured Cells DNA Purification Kit (Cat#AS1620) on the Promega Maxwell 48 RSC instrument (Madison, WI). Genomic DNA was sheared using the Covaris ME220 Focused-ultrasonicator (Woburn, MA). Indexed libraries were prepared using the Tecan Ovation Ultralow System V2 Kit (San Carlos, CA) and PerkinElmer Zephyr G3 NGS Workstation (Waltham, MA). Libraries were analyzed using the Standard Sensitivity NGS Fragment Analysis Kit and Advanced Analytical Fragment Analyzer System (Ankeny, IA). WGS was performed using either the MiSeq Reagent V2 500 cycle Kit and Illumina MiSeq System or the P1 600 Cycle NextSeq reagent kit and the Illumina NextSeq System (San Diego, CA), generating 2×250 or 2×300 paired-end reads, respectively. Sequence data were analyzed for quality, sequence type (ST), and AR genes using the bioinformatics pipeline PHoeNIx v2.1.1 [28]. Sequences are uploaded to BioProject accession no. PRJNA1280115.

### Wastewater samples and clinical testing correlation

De-identified clinical testing (all facilities) and colonization screening results (Facility C only) during the WWS timeframes were provided by the Georgia Department of Public Health (GA DPH) if available. Clinical and screening results were compared to isolates from the closest wastewater testing dates within a two-week period. For any matching clinical/colonization and WWS isolates identified within the two-week window, isolates were compared using PCR or bioinformatically using SNVPhyl [29].

### Workflow comparison and notable agar findings

Exact McNemar’s test (p-value ≤0.05) was used to compare the detection of *bla*_KPC_, *bla*_NDM_, *bla*_VIM_, *bla*_OXA-48-like_, and *bla*_IMP_ from the ‘enrichment-to-isolate’ and ‘enrichment-to-PCR’ workflows using Microsoft Excel (version 2408, Microsoft Corporation, Redmond, Washington). Notable agar findings (e.g., isolate coloration) discordant with the instructions for use (IFUs) were documented.

## Results

### Facilities and wastewater sampling

Facility A reported a mean resident census of 169.8 (range: 157-179), and 23 weekly wastewater samples were collected. Facility B reported a mean resident census of 79.7 (range: 74-87), and 25 wastewater samples were collected. Facility C reported a mean resident census of 92.4 (range: 88-99), and 25 wastewater samples were collected.

### Wastewater Enrichment and E. coli Detection Using Colilert-18 Assay

Colilert-18 assay estimated an *E. coli* MPN median of 4.9 log_10_ MPN/100mL (±6.3) for Facility A, 4.7 log_10_ MPN/100mL (±6.7) for Facility B, and 6.2 log_10_ MPN/100mL (±1.7) for Facility C.

### Culture-based Detection of Carbapenem-Resistant Bacteria

*Enrichment-to-isolate*: Table 1 and Figure 2 detail CR bacterial isolates, separated by clinical and environmental/opportunistic relevance, from the SNFs’ wastewater per the ‘enrichment-to-isolate’ workflow. Similar genera were identified in wastewater from all three facilities, including four clinically relevant complexes/species - *Acinetobacter baumannii* complex, *Enterobacter cloacae* complex, *Enterococcus faecium*, and *Stenotrophomonas maltophilia*, and three environmental/opportunistic complexes/species - *Comamonas testosteroni*, *Pandoraea apista*, and *Pseudomonas putida* group (Table 1). Each SNF had one clinically relevant complex/species unique to that facility: *Aeromonas hydrophila* (Facility A), *Burkholderia cepacia complex* (Facility B), and *Serratia marcescens* (Facility C). *Citrobacter freundii*, *Escherichia coli*, and *Klebsiella oxytoca* were detected in Facilities A and C, and *Klebsiella pneumoniae* and *Proteus* spp. were detected in Facilities B and C.

**Figure 2.**
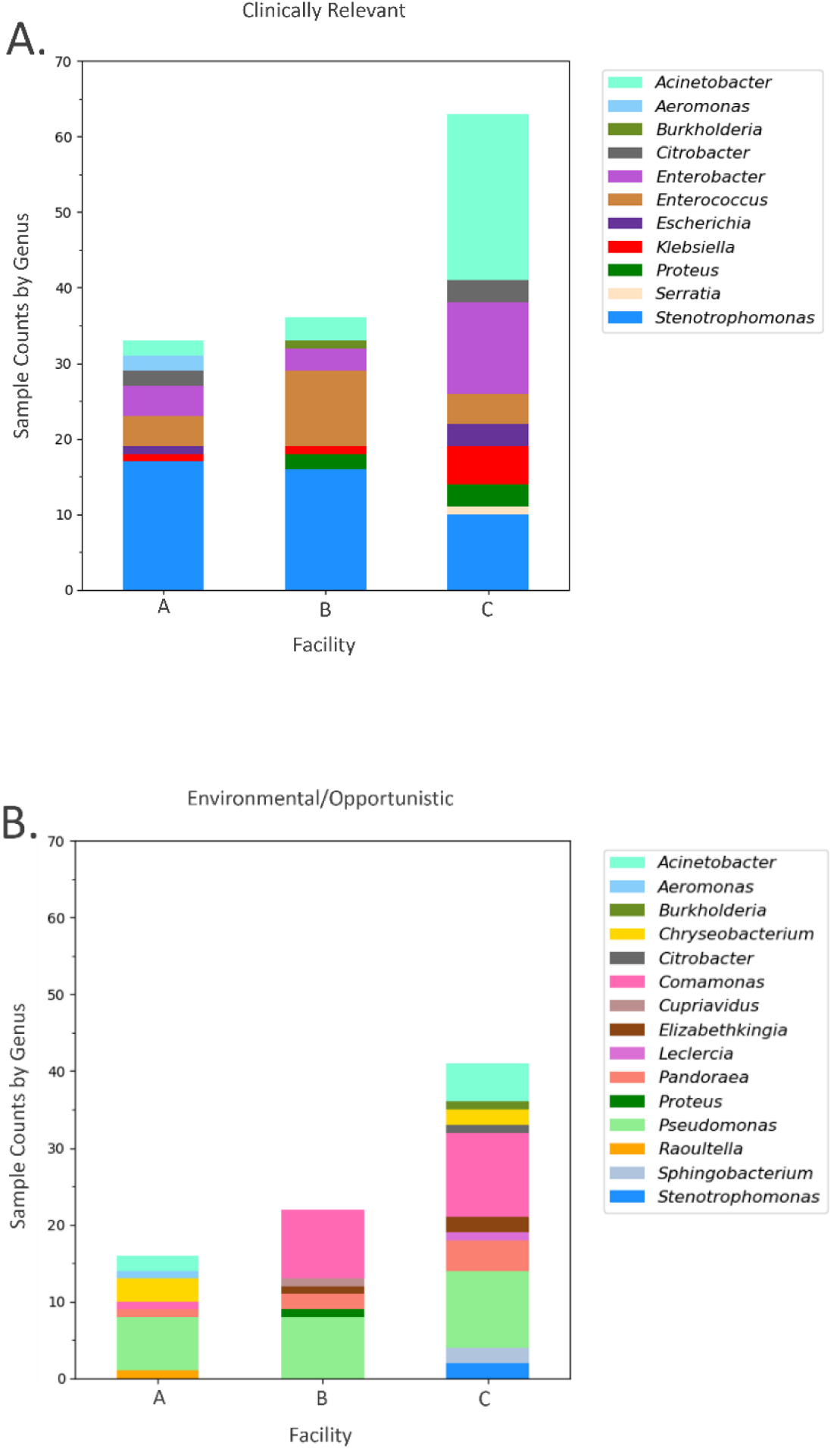
Carbapenem-resistant bacteria isolated from wastewater samples from three skilled nursing facilities (Georgia, USA; 2022). The colors reflect genera detected by MALDI-TOF identification of antimicrobial-resistant isolates collected from Facility A (n=23 samples), Facility B (n=25), and Facility C (n=25); (2A) Clinically relevant and (2B) Environmental/opportunistic carbapenem-resistant bacteria in healthcare settings.^Ŧ^ See Supplemental for species level details. ^Ŧ^ Acinetobacter, Aeromonas, Burkholderia, Citrobacter, Proteus, and Stenotrophomonas contain both clinically relevant and environmental/opportunistic organisms (2A) Clinically relevant: Acinetobacter baumannii complex; Aeromonas hydrophila; Burkholderia cepacia complex; Citrobacter freundii; Proteus vulgaris group; Stenotrophomonas maltophilia (2B) Environmental/Opportunistic: Acinetobacter: baumannii complex, baylyi, radioresistens, ursingii; Aeromonas caviae; Burkholderia cepacia complex; Citrobacter braakii; Proteus vulgaris group; Stenotrophomonas acidaminiphila

**Table 1:**
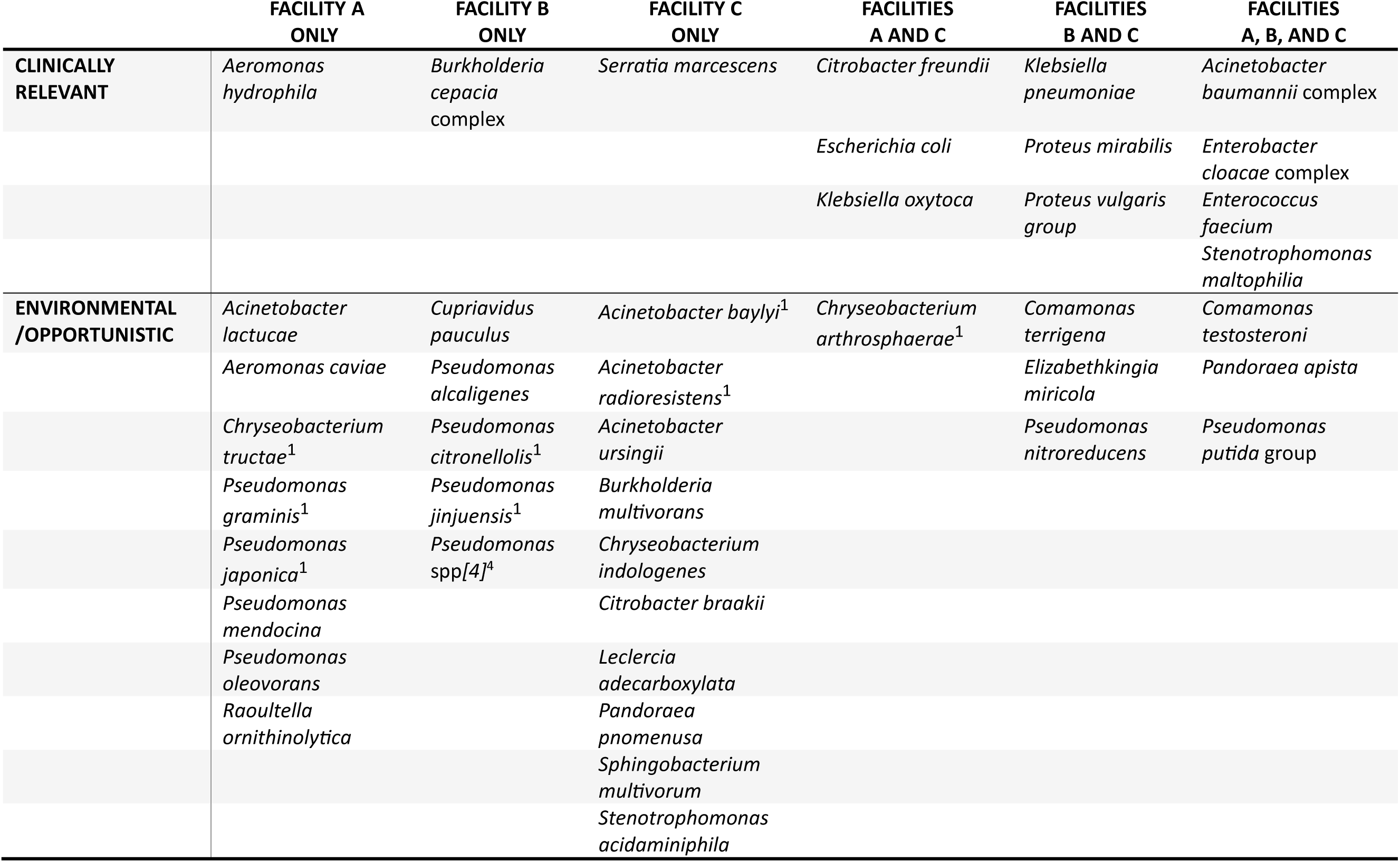
Carbapenem-resistant bacterial community as identified by MALDI-TOF after growth on selective media from three skilled nursing facilities’ wastewater^2^ (Georgia, USA; 2022). All bacteria listed had a confident species identification (i.e., confidence score: 2.000-3.000), unless otherwise noted^3^. ^1^Organisms with a MADLI-TOF score of 1.700 – 1.999 are included (confident genus identification, possible species identification ovided by MALDI-TOF output); ^2^ No shared species between Facility A and B; ^3^ https://microbenet.cdc.gov/; ^4^ Unclassified *eudomonas* species

The genera most frequently isolated from all three facilities’ wastewater samples were *Stenotrophomonas* spp. (Fig. 2a) and *Pseudomonas* spp. (Fig. 2b). Clinically relevant bacteria detected in at least 25% of samples were *Enterococcus* spp. from Facility B, and *Acinetobacter* spp. and *Enterobacter* spp. from Facility C. The environmental/opportunistic relevant bacteria detected in at least 25% of samples in each facility were *Comamonas* spp. in Facilities B and C. Facility A did not have any other genus in at least 25% of the samples beyond *Stenotrophomonas* spp. and *Pseudomonas* spp.

*Enrichment-to-PCR*: Four Facility A pooled lysate wastewater samples were PCR-positive for *bla*_KPC_ (Supplemental Table 1). There were no PCR-positive results for *bla*_NDM_, *bla*_OXA-48-like_, *bla*_VIM_, or *bla*_IMP_ in Facility A, and no PCR-positive results in Facility B. Seven Facility C pooled lysate samples were PCR-positive for *bla*_KPC_ and one for *bla*_OXA-48-like_ (Supplemental Table 1). There were no PCR-positive results for *bla*_NDM_, *bla*_VIM_, or *bla*_IMP_ in Facility C.

### Molecular-based Detection of Carbapenemase Genes

*PCR workflow*: Nine Facility A isolates from 8 (34.8%) of 23 pooled wastewater samples were PCR-positive for *bla*_KPC_ (Table 2). *bla*_KPC_-positive *E. hormaechei* was only found in Facility A and was the most frequently isolated PCR-positive species found in Facility A (five isolates from four composite samples across six months). *C. freundii* was the next most identified *bla*_KPC_-positive bacteria in Facility A, found in two composite samples in the same month. There were no PCR-positive isolates from Facility B. Fourteen Facility C isolates from 10 (40.0%) of 25 total pooled wastewater samples were PCR-positive for *bla*_KPC_. The two most identified *bla*_KPC_-positive bacteria in Facility C were *C. freundii* and *E. roggenkampii* (four isolates each).

**Table 2.** Wastewater isolate characterization. Facility A and C wastewater isolates identified by MADLI-TOF after growth on selective agar (Georgia, USA; 2022); carbapenemase (CP) genes by PCR and whole genome sequencing (WGS); additional beta-lactamase resistance genes (beyond the big 5 CP genes) identified by WGS. Clinically relevant isolates are in bold. Facility B did not have any isolates with CP genes detected. (Sequences are uploaded to BioProject accession no. PRJNA1280115).

| FACILITY-SAMPLE # | ORGANISM | CP GENE BY PCR | CP gene by WGS | ADDITIONAL BETA-LACTAMASE GENES BY WGS |
| --- | --- | --- | --- | --- |
| A-21 | *Citrobacter freundii* | *bla*<sub>KPC</sub> | *bla*<sub>KPC-4</sub> | *bla*<sub>CARB-2</sub>; *bla*<sub>CMY-116</sub>; *bla*<sub>TEM-1A</sub> |
| A-23 | *Citrobacter freundii* | *bla*<sub>KPC</sub> | *bla*<sub>KPC-4</sub> | *bla*<sub>CMY-116</sub>; *bla*<sub>TEM-1A</sub> |
| A-2 | *Enterobacter hormaechei* | *bla*<sub>KPC</sub> | *bla*<sub>KPC-4</sub> | *bla*<sub>ACT-46</sub>; *bla*<sub>TEM-1A</sub> |
| A-2 | *Enterobacter hormaechei* | *bla*<sub>KPC</sub> | *bla*<sub>KPC-3</sub> | *bla*<sub>ACT-55</sub> |
| A-6 | *Enterobacter hormaechei* | *bla*<sub>KPC</sub> | *bla*<sub>KPC-3</sub> | *bla*<sub>ACT-55</sub> |
| A-14 | *Enterobacter hormaechei* | *bla*<sub>KPC</sub> | *bla*<sub>KPC-4</sub> | *bla*<sub>OXA-1</sub>; *bla*<sub>ACT-46</sub>; *bla*<sub>TEM-1A</sub> |
| A-20 | *Enterobacter hormaechei* | *bla*<sub>KPC</sub> | *bla*<sub>KPC-4</sub> | *bla*<sub>OXA-1</sub>; *bla*<sub>ACT-46</sub>; *bla*<sub>TEM-1A</sub> |
| A-8 | *Klebsiella oxytoca* | *bla*<sub>KPC</sub> | *bla*<sub>KPC-3</sub> | *bla*<sub>OXA-9</sub>; *bla*<sub>OXY-6-4</sub>; *bla*<sub>TEM-1A</sub> |
| A-10 | *Pseudomonas mendocina* | *bla*<sub>KPC</sub> | *bla*<sub>KPC-2</sub> |  |
| C-17 | *Acinetobacter baumannii* | *bla*<sub>OXA-23</sub> | *bla*<sub>OXA-23</sub> | *bla*<sub>OXA-66</sub>; *Mbl*; *Zn-dependent hydrolase*; *bla*<sub>ADC-30</sub> |
| C-19 | *Acinetobacter baumannii* | *bla*<sub>OXA-23</sub> | *bla*<sub>OXA-23</sub> | *bla*<sub>OXA-66</sub>; *Mbl*; *Zn-dependent hydrolase*; *bla*<sub>ADC-30</sub> |
| C-12 | *Citrobacter braakii* | *bla*<sub>KPC</sub> | *bla*<sub>KPC-3</sub> | *bla*<sub>CMY-34</sub> |
| C-12 | *Citrobacter freundii* | *bla*<sub>KPC</sub> | *bla*<sub>KPC-3</sub> | *bla*<sub>CMY-34</sub> |
| C-17 | *Citrobacter freundii* | *bla*<sub>KPC</sub> | *bla*<sub>KPC-3</sub> | *bla*<sub>CMY-34</sub> |
| C-18 | *Citrobacter freundii* | *bla*<sub>KPC</sub> | *bla*<sub>KPC-3</sub> | *bla*<sub>CMY-34</sub> |
| C-18 | *Citrobacter freundii* | *bla*<sub>KPC</sub> | *bla*<sub>KPC-3</sub> | *bla*<sub>CMY-34</sub> |
| C-19 | *Enterobacter ludwigii* | *bla*<sub>KPC</sub> | *bla*<sub>KPC-3</sub> | *bla*<sub>ACT-12</sub> |
| C-22 | *Enterobacter ludwigii* | *bla*<sub>KPC</sub> | *bla*<sub>KPC-3</sub> | *bla*<sub>ACT-12</sub> |
| C-3 | *Enterobacter roggenkampii* | *bla*<sub>KPC</sub> | *bla*<sub>KPC-3</sub> | *bla*<sub>MIR-16</sub> |
| **C-11** | ***Enterobacter roggenkampii*** | *bla*<sub>KPC</sub> | *bla*<sub>KPC-3</sub> | *bla*<sub>MIR-16</sub> |
| **C-12** | ***Enterobacter roggenkampii*** | *bla*<sub>KPC</sub> | *bla*<sub>KPC-3</sub> | *bla*<sub>MIR-16</sub> |
| **C-15** | ***Enterobacter roggenkampii*** | *bla*<sub>KPC</sub> | *bla*<sub>KPC-3</sub> | *bla*<sub>MIR-16</sub> |
| **C-4** | ***Klebsiella oxytoca*** | *bla*<sub>KPC</sub> | *bla*<sub>KPC-3</sub> | *bla*<sub>OXY-5-1</sub> |
| **C-12** | ***Klebsiella oxytoca*** | *bla*<sub>KPC</sub> | *bla*<sub>KPC-3</sub> | *bla*<sub>OXY-5-1</sub> |
| **C-1** | ***Leclercia adecarboxylata*** | *bla*<sub>KPC</sub> | *bla*<sub>KPC-3</sub> | *bla*<sub>SHV-12</sub>; *bla*<sub>TEM-1</sub> |

*Whole genome sequencing workflow:* WGS was performed on 88.4% (30 of 34) of Facility A isolates, 90.4% (47 of 52) of Facility B, and 85.4% (105 of 123) of Facility C. No discrepancies were found between the PCR and WGS results (100% concordance, 182 of 182). The bacteria identified by MALDI-TOF with negative PCR results for the five targeted CR genes but detection of other beta-lactamase genes at the three SNFs are detailed in Supplemental Table 2. Of note, 41.0% (21 of 51) of PCR-negative isolates from Facility C with other beta-lactamase genes identified by WGS (i.e., those not targeted by the PCR assays) were *A. baumannii* (see Supplemental Table 2).

### Wastewater samples and clinical testing correlation

At Facility A, KPC-positive *K. oxytoca* was isolated from a clinical specimen (urine) on April 18, 2022, and detected in only one wastewater sample (April 5, 2022). Sequencing data revealed these isolates are highly related (differing by one high-quality single-nucleotide variant, across a core genome of 99.3%) and carried the same KPC variant, *bla*_KPC-3_.

At Facility C, a *bla*_OXA-23_-positive *A. baumannii* was isolated from a resident’s clinical specimen (urine) on November 2. Two of three wastewater *A. baumannii* isolates from the proximal three sampling dates (October 25, November 1, and November 8, 2022) were positive for *bla*_OXA-23_ by PCR and WGS (October 25 and November 8, 2022). Additionally, WGS revealed six other *bla*_OXA-23_ *A. baumannii*-positive wastewater samples during this study period before the positive clinical specimen; however, resident clinical specimen sequences were not available to align with the wastewater sample sequences.

Screening for CROs was not conducted at Facility B.

### Workflow comparison and notable agar findings

*Agars and clinically relevant bacteria: Acinetobacter baumannii* complex, *Citrobacter freundii*, *Enterobacter cloacae* complex, *Enterococcus faecium*, and *Stenotrophomonas maltophilia* were the species/complexes most frequently detected of the 13 carbapenem-resistant clinically relevant complexes/groups from the SNF wastewater samples (Fig. 3a). mSC agar captured 23 detections of CRE overall. In the only instance where mSC did not detect an organism that another agar detected (AC agar, *B. cepacia*), the agar was not developed to support this genus. The non-CRE species/complexes detected in the wastewater samples by mSC were *Acinetobacter* spp., *Aeromonas* spp., *Enterococcus* spp., and S*tenotrophomonas* spp. MAC+erta and MAC+mero agar missed three and five clinically relevant bacteria, respectively, as compared to mSC (Fig. 3a). The AC agar, specific for the detection of *Acinetobacter* spp., missed 13 detections that mSC captured, but had 40 detections of *S. maltophilia*. The PC agar isolated *Pseudomonas* spp. in two samples, while other agars identified the bacteria in 20 samples (data not shown).

**Figure 3.**
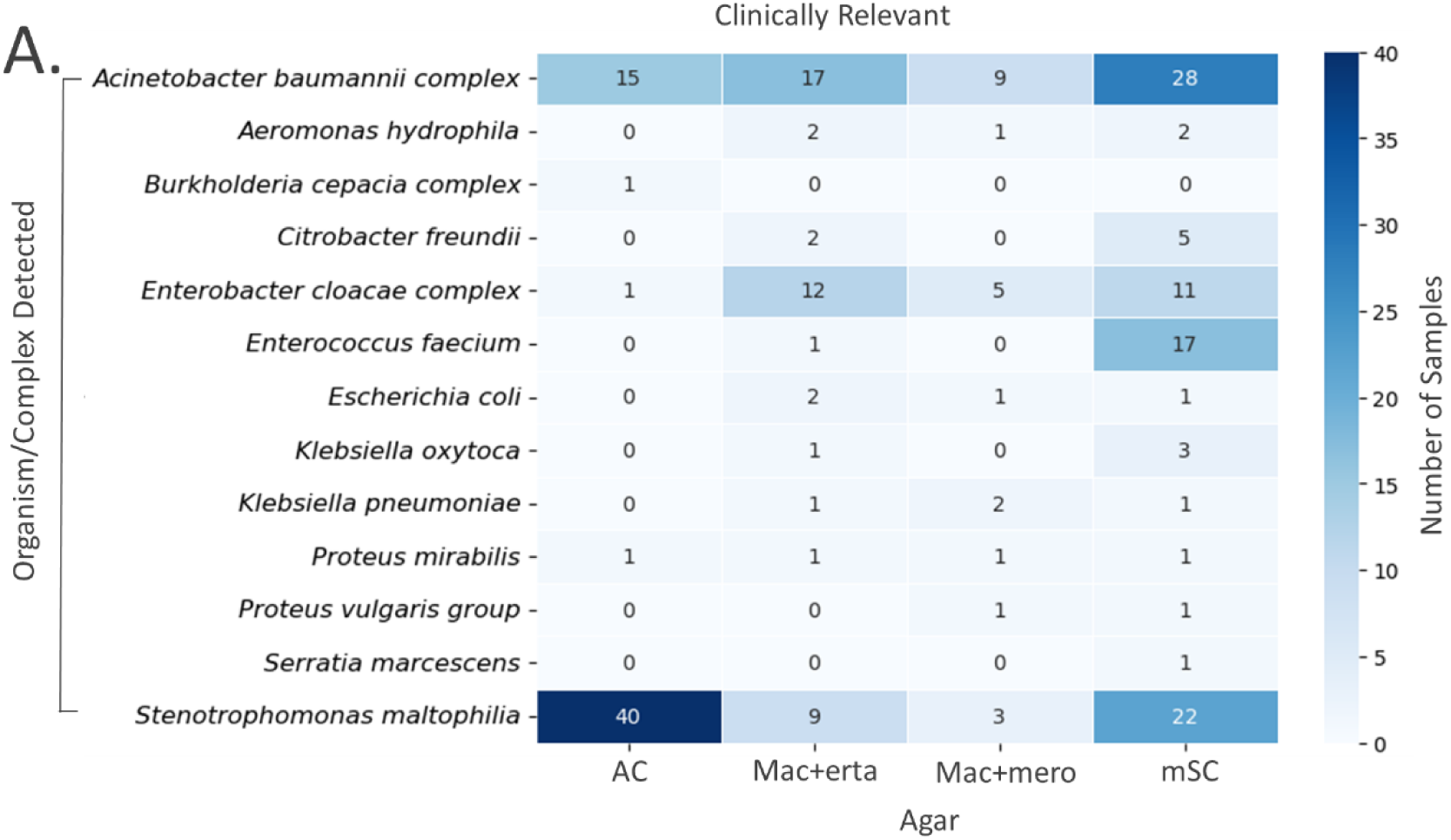
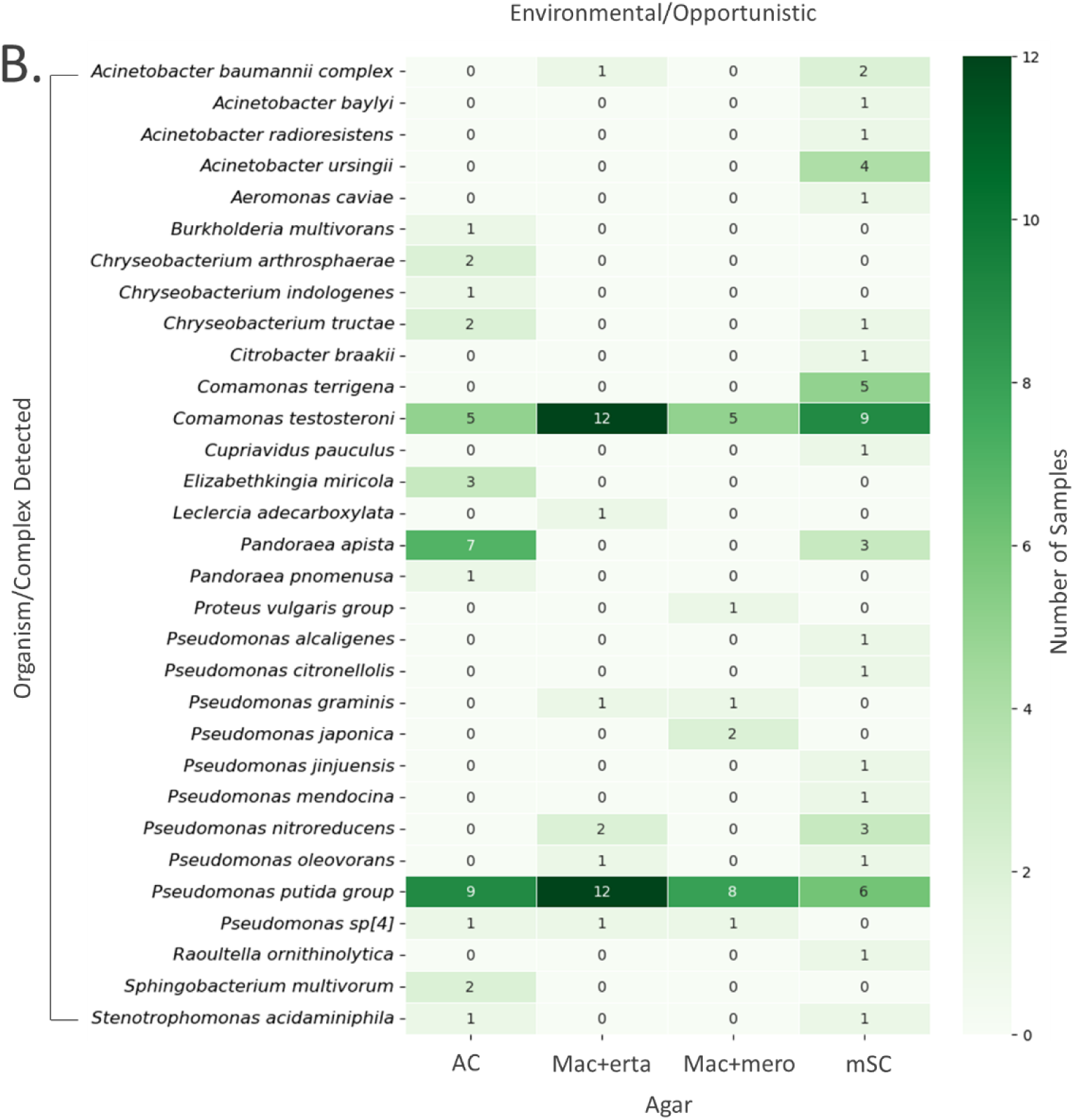
Heatmap of the organisms cultured from three skilled nursing facilities’ wastewater (Georgia, USA; 2022) on selective agars, identified using MALDI-TOF. (3A) Clinically relevant and (3B) Environmental/opportunistic carbapenem-resistant bacteria in healthcare settings (AC = CHROMagar Acinetobacter + MDR Supplement. MAC+erta = MacConkey II agar + ertapenem (10µg disk). MAC+mero = MacConkey II agar + meropenem (10µg disk). mSC = CHROMagar mSuperCARBA, where agar types are not mutually exclusive. Total number of composite samples collected, n = 73.^Ŧ^ ^Ŧ^ *Acinetobacter baumannii* complex and *Proteus vulgaris* group contain both clinically relevant and environmental/opportunistic organisms. Species were counted individually but reported within their respective complex or group according to reporting guidelines.

*Agars and environmental/opportunistic bacteria*: *Comamonas terrigena*, *Comamonas testosteroni*, *Pandoraea apista*, *Pseudomonas nitroreducens*, and *Pseudomonas putida* group were the most frequently detected species/complexes of the 31 carbapenem-resistant environmental/opportunistic species/complexes from the SNF wastewater samples (Fig. 3b). Detections of other environmental/opportunistic species/complexes were typically less than five occurrences of the 73 total wastewater samples. There were 31 instances in which a given species was detected from a single agar (Fig. 3b).

*Workflow comparison*: A total of 67 sample pairs were available for comparison of the enrichment-to-isolate (gold standard; using mSC agar only) and enrichment-to-PCR workflows (Table 3). Concordance between the methods for detecting or not detecting any CR-gene occurred in 57 (85%) of 67 samples, noting that 48 were paired negative results (both isolate and pooled lysate) and nine were paired positive results (both isolate and pooled lysate). There was no significant difference between the two workflows for the detection of any targeted CR-gene (McNemar’s test; p-value = 0.21). The sensitivity of the enrichment-to-PCR workflow was 56.3%, and specificity was 94.1%.

**Table 3.**
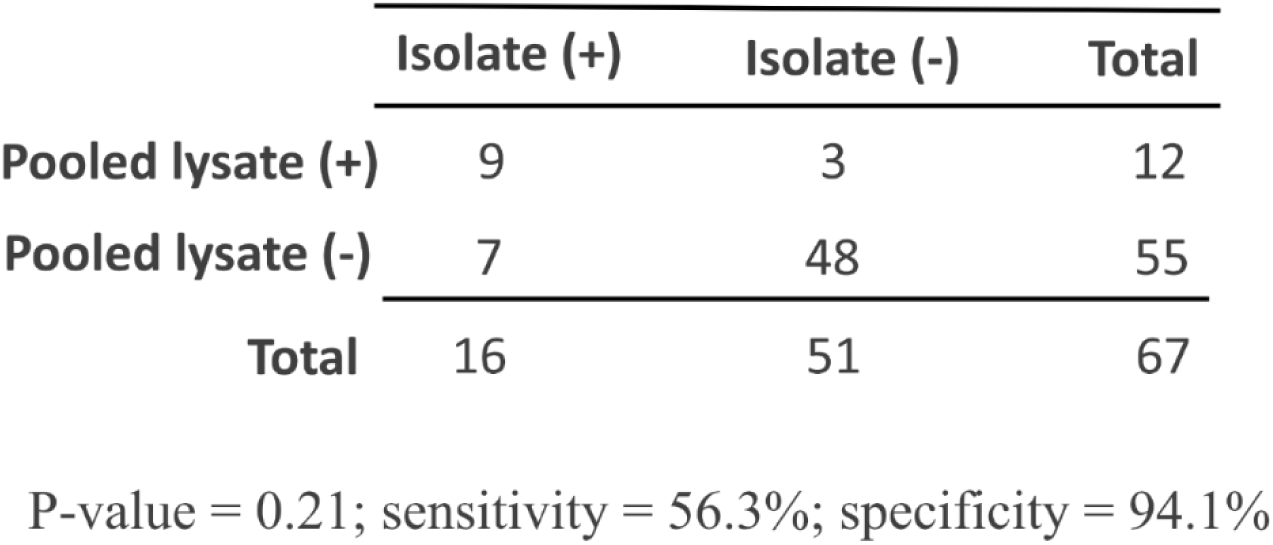
Comparison of the ‘enrichment-to-isolate’ (Isolate(+), Isolate(−); mSC agar only) and ‘enrichment-to-PCR’ (Pooled lysate(+), Pooled lysate(−)) workflows, sensitivity, and specificity for the detection of carbapenem-resistant genes (*bla*_KPC_, *bla*_NDM_, *bla*_VIM_, *bla*_OXA-48-like_, and *bla*_VIM_) from wastewater samples in three skilled nursing facilities (Georgia, USA; 2022), as determined by McNemar’s test, where statistical significance is set at P-value≤0.05. This analysis includes 67 of the 73 total pooled samples that were available for comparison.

|  | Isolate (+) | Isolate (-) | Total |
| --- | --- | --- | --- |
| **Pooled lysate (+)** | 9 | 3 | 12 |
| **Pooled lysate (-)** | 7 | 48 | 55 |
| **Total** | 16 | 51 | 67 |

P-value = 0.21; sensitivity = 56.3%; specificity = 94.1%

All samples with a PCR-positive result from any agar (n=21) were compared for the two workflows (Supplemental Table 1). Both workflows detected *bla*_KPC_ in nine samples. However, the enrichment-to-isolate workflow detected *bla*_KPC_ in nine additional samples that enrichment-to-PCR did not, and enrichment-to-PCR detected two *bla*_KPC_ and one *bla_OXA_*_-48-like_ in three samples that enrichment-to-isolate did not. The enrichment-to-isolate workflow detected 85.7% (18 of 21), and the enrichment-to-PCR workflow detected 57.1% (12 of 21).

## Discussion

WWS in healthcare settings for AR is in the early stages of development and applications. This study successfully demonstrated the feasibility of applying existing methodologies to SNF wastewater samples for detecting and surveying CR bacteria and genes. mSC agar outperformed the other selective agars in detecting the targeted CR bacteria. All clinically relevant Enterobacteriaceae identified on other agar types were also identified on mSC, and mSC was the most effective at isolating *Acinetobacter* spp. and *Pseudomonas* spp. from SNF wastewater samples. Most of the PCR-positive CR isolates identified were from clinically relevant bacteria. These two findings highlight that mSC and the molecular methods, including both PCR and WGS, were effective in detecting clinically important targets from SNF wastewater.

This study used the Colilert-18 assay as a simple processing method to enrich for *E. coli* and coliform bacteria for downstream AR detection applications, similar to previous international efforts [20, 30]. The PCR assays, which are used in the AR Lab Network for clinical isolates, detected carbapenemase genes and aligned with the WGS data. Additionally, international studies that screened for CRE in healthcare setting wastewater found many of the same genera and species (i.e., *Escherichia* spp., *Klebsiella* spp., *Enterobacter* spp.), though the prevalence of the main CR genes varied according to the region [31, 32]. *bla*_KPC_ was found to be the dominant carbapenemase gene, which is not unexpected given it is the most common carbapenemase gene in the United States [33]. Similarly, a review focused on international countries regarding antimicrobial-resistant bacteria and AR gene detection in hospital/hospital-adjacent area wastewater described seven studies that detected *bla*_NDM_ and one study that detected four of the five main CR genes (*bla*_KPC_, *bla*_NDM_, *bla*_OXA-48_, *bla*_VIM_), demonstrating wastewater detection of genes circulating in each country [34].

However, the applied workflows have room for optimization to be more selective for CR bacteria in wastewater samples (e.g., integrating antibiotics to the Colilert-18 assay [35, 36] and switching from NaOH thermal lysis to a commercial kit-based extraction method). Although the ability to pre-screen pooled lysate samples from wastewater using PCR before plating would be ideal, the enrichment-to-PCR workflow (pooled lysate samples), as implemented here, had low sensitivity (56.3%) compared to the enrichment-to-isolate workflow. Specific to the enrichment-to-isolate workflow, it is possible that some bacteria carrying carbapenemase genes were not captured by isolate analysis. This may be due to several limitations within this study, including choosing representative unique colony morphologies from each selective agar, which may not capture all genera or species present, and not saving *Stenotrophomonas* or *Aeromonas* isolates for downstream analysis due to their intrinsic resistance to beta-lactams [24]. A notable challenge of using mSC to culture wastewater samples, a complex matrix, is some bacteria may differ from the expected morphology. Additional notable findings detailing discrepancies in wastewater isolates on the tested agars, per the IFUs, are discussed in detail (Supplemental Materials). Future studies should continue to integrate culture and molecular approaches, as others have also reported value in validating sequencing data with culture confirmation [15]. Furthermore, PCR-based methods are limited in the number of AR gene targets, which must also be selected *a priori*, meaning novel AR genes would go undetected. Sequencing approaches could help address these limitations and therefore support efforts to better understand silent circulation of new or emerging resistance and potentially slow regional AR transmission with earlier and more frequent surveillance as compared to intermittent patient screening practices (i.e., admission, prevention).

One of the current limitations of WWS as a public health tool is the direct correlation of findings in wastewater to clinical incidence. One consideration, as applied here, is to integrate and adapt assays used by public health departments and the AR Lab Network to provide strength in direct comparisons and data interpretation. While we provided two single scenarios for proof-of-concept linkage of wastewater sample data with retrospectively collected clinical testing results from two facilities, it was beyond the scope in 2022 to actively collect or ask the facility for AR clinical data during the COVID-19 pandemic. Additionally, the challenge of differentiating relevant signals for infection control action from baseline or background antimicrobial-resistant signals is notable. More development is needed to align facility patient testing data with healthcare facility WWS before this approach is feasible for proactive infection prevention and control. There remains significant value in pursuing systematic, robust healthcare facility WWS efforts to aid in early detection of emerging antimicrobial-resistant bacteria and AR genes.

## Data availability

Sequencing data generated from this manuscript are available at BioProject PRJNA1280115.

## Acknowledgements

This project was supported in part by an appointment to the Research Participation Program at the Centers for Disease Control and Prevention administered by the Oak Ridge Institute for Science and Education through an interagency agreement between the U.S. Department of Energy and the Centers for Disease Control and Prevention. We extend our gratitude to the Georgia Department of Public Health for their guidance and the skilled nursing facilities that participated. We would also like to thank Alyssa Kent for her bioinformatic support and Thomas Ewing, Davina Campbell, Sarah Sabour, and Cynthia Longo for their technical guidance.

## Author contributions

ACS, LF, SL: Conceived of the presented idea; ACS, SL: Wrote manuscript; SL: Conducted laboratory processes for samples (culture, MALDI, sequencing); SB: Conducted laboratory processes for culture, MALDI-TOF, and PCR, contributed to and reviewed manuscript; IT: Conducted laboratory processes for culture, MALDI-TOF, and PCR, contributed to and reviewed manuscript; LTB: Conducted laboratory processes for culture, MALDI-TOF, and PCR, contributed to and reviewed manuscript; AKL: Contributed to facility partnership, supported laboratory processing, contributed to and reviewed manuscript; TM: Created figures 2 and three, contributed to and reviewed manuscript; FW: contributed to facility partnership and clinical testing alignment, contributed to and reviewed manuscript; ALH: Sequencing expertise and guidance, contributed to and reviewed manuscript and associated poster/presentations; MK: Clinical antimicrobial resistance expertise and guidance, contributed to and reviewed manuscript and associated poster/presentations; MW: Contributed to and reviewed manuscript and associated poster/presentations; LF: Clinical antimicrobial resistance expertise and guidance, contributed to and reviewed manuscript and associated poster/presentations; ACS: Wastewater surveillance expertise and guidance.

## Footnote Page

§See e.g., 45 C.F.R. part 46, 21 C.F.R. part 56; 42 U.S.C. §241(d); 5 U.S.C. §552a; 44 U.S.C. §3501 et seq.

## Meetings where the information has previously been presented

(Poster) Susanna Lenz MS, Subhasish Biswas BS, Ivy Terry MPH, Lisa Tran Basha BS, Amanda Lyons MS, MPH, Florence Whitehill DVM, MPH, Tapati Mazumdar PhD, MPH, Alison Laufer Halpin PhD, Lauren Franco PhD, D(ABMM), Maria Karlsson PhD, Margaret Williams PhD, Angela Coulliette-Salmond MS, PhD. “Carbapenem-Resistant Organism Detection in Skilled Nursing Facility Wastewater.” Microbes in Wastewater: Antibiotic Resistance, Public Health, and Climate Change. January 16-17, 2025. Newport Beach, California.

