## Supplemental Materials for "Detection of Carbapenem-Resistant Bacteria in Skilled Nursing Facility Wastewater"


- Pg. 1 - Supplemental Table 1. PCR-positive results of pooled lysate samples and carbapenem-resistant (CR) bacterial isolates from Facilities A and C (no samples from facility B were PCR-positive)
- Pg. 2 - Supplemental Table 2. PCR-Negative Organisms with Beta-lactamase Genes
- Pg. 4 - Notable Findings


Supplemental Table 1. PCR-positive results of pooled lysate samples from the enrichment-to-PCR workflow and carbapenem-resistant (CR) bacterial isolates from the enrichment-to-isolate workflow (Georgia, USA; 2022); Facilities A and C captured on the selective agars compared (no samples from Facility B were PCR-positive).

| FACILITY-SAMPLE # | POOLED LYSATE | ISOLATE | POOLED LYSATE | ISOLATE |
|---|---|---|---|---|
| | **KPC** | **KPC** | **OXA-48** | **OXA-48** |
| **A-2** | + | + | - | - |
| **A-6** | + | + | - | - |
| **A-8** | - | + | - | - |
| **A-10** | - | + | - | - |
| **A-14** | - | + | - | - |
| **A-19** | + | - | - | - |
| **A-20** | + | + | - | - |
| **A-21** | - | + | - | - |
| **A-23** | - | + | - | - |
| **C-1** | - | + | - | - |
| **C-2** | + | - | - | - |
| **C-3** | + | + | - | - |
| **C-4** | + | + | - | - |
| **C-10** | - | - | + | - |
| **C-11** | - | + | - | - |
| **C-12** | + | + | - | - |
| **C-15** | - | + | - | - |
| **C-17** | + | + | - | - |
| **C-18** | + | + | - | - |
| **C-19** | - | + | - | - |
| **C-22** | + | + | - | - |

Supplemental Table 2. Wastewater isolate characterization. Facility A, B, and C wastewater isolates (Georgia, USA; 2022) identified by MADLI-TOF after growth on selective agar, and beta-lactamase genes identified by whole genome sequencing of isolates PCR-negative for targeted carbapenemase genes. Clinically relevant isolates are in bold.

| FACILITY-SAMPLE # | ORGANISM | ADDITIONAL BETA-LACTAMASE GENES |
|---|---|---|
| **A-20** | ***Acinetobacter baumannii*** | $bla_{OXA-23}$; $bla_{OXA-66}$; *Mbl*; *Zn-dependent hydrolase*; $bla_{ADC-30}$ |
| **A-23** | ***Acinetobacter baumannii*** | $bla_{OXA-51}$; $bla_{A2}$; *Zn-dependent hydrolase*; $bla_{ADC-165}$ |
| **A-3** | *Acinetobacter lactucae* | $bla_{OXA-500}$; $bla_{ADC-18}$ |
| **A-20** | *Acinetobacter lactucae* | $bla_{OXA-819}$ |
| **A-6** | *Chryseobacterium tructae* | $bla_{IND-8}$ |
| **A-23** | *Chryseobacterium tructae* | $bla_{IND-8}$ |
| **A-8** | ***Escherichia coli*** | *AmpC1 Ecoli*; *Penicillin Binding Protein Ecoli*; *ampH Ecoli*; $bla_{CMY-2}$; $bla_{EC-19}$ |
| **A-18** | *Pandorea apista* | $bla_{OXA-153}$ |
| **A-18** | *Raoultella ornithinolytica* | $bla_{ORN-1}$ |
| **B-20** | ***Acinetobacter baumannii*** | $bla_{OXA-113}$; $bla_{OXA-398}$; *Zn-dependent hydrolase*; $bla_{ADC-222}$ |
| **B-22** | ***Acinetobacter baumannii*** | $bla_{OXA-113}$; $bla_{OXA-398}$; *Zn-dependent hydrolase*; $bla_{ADC-222}$ |
| **B-21** | ***Acinetobacter nosocomialis*** | *Mbl*; $bla_{ADC-130}$ |
| **B-22** | ***Acinetobacter nosocomialis*** | $bla_{ADC-289}$ |
| **B-18** | *Elizabethkingia miricola* | $bla_{B-27}$; $bla_{GOB-37}$ |
| **B-22** | ***Enterobacter hormaechei*** | $bla_{ACT-40}$ |
| **B-3** | ***Enterobacter roggenkampii*** | $bla_{MIR-22}$ |
| **B-9** | ***Enterobacter roggenkampii*** | $bla_{MIR-15}$ |
| **B-10** | ***Enterobacter roggenkampii*** | $bla_{MIR-5}$ |
| **B-13** | ***Enterobacter roggenkampii*** | $bla_{MIR-15}$ |
| **B-22** | ***Enterobacter roggenkampii*** | $bla_{MIR-15}$ |
| **B-10** | ***Klebsiella pneumoniae*** | *ampH*; $bla_{SHV-168}$ |
| **B-1** | ***Klebsiella variicola*** | *ampH*; $bla_{LEN-11}$ |
| **B-4** | *Pandoraea apista* | $bla_{OXA-153}$ |
| **B-19** | *Pandoraea apista* | $bla_{OXA-153}$ |
| **C-2** | ***Acinetobacter baumannii*** | $bla_{OXA-68}$; *Mbl*; *Zn-dependent hydrolase*; $bla_{ADC-76}$ |
| **C-3** | ***Acinetobacter baumannii*** | $bla_{OXA-23}$; $bla_{OXA-66}$; *Mbl*; *Zn-dependent hydrolase*; $bla_{ADC-30}$ |
| **C-4** | ***Acinetobacter baumannii*** | $bla_{OXA-23}$; $bla_{OXA-66}$; *Mbl*; *Zn-dependent hydrolase*; $bla_{ADC-30}$ |
| **C-5** | ***Acinetobacter baumannii*** | $bla_{OXA-23}$; $bla_{OXA-66}$; *Mbl*; *Zn-dependent hydrolase*; $bla_{ADC-30}$ |
| **C-6** | ***Acinetobacter baumannii*** | $bla_{OXA-23}$; $bla_{OXA-66}$; *Mbl*; *Zn-dependent hydrolase*; $bla_{ADC-30}$ |
| **C-7** | ***Acinetobacter baumannii*** | $bla_{OXA-23}$; $bla_{OXA-66}$; *Mbl*; *Zn-dependent hydrolase*; $bla_{ADC-30}$ |
| **C-8** | ***Acinetobacter baumannii*** | $bla_{OXA-68}$; *Mbl*; *Zn-dependent hydrolase*; $bla_{ADC-277}$; $bla_{ADC-76}$ |
| **C-9** | ***Acinetobacter baumannii*** | $bla_{OXA-117}$; *Mbl*; *Zn-dependent hydrolase*; $bla_{ADC-238}$ |
| **C-10** | ***Acinetobacter baumannii*** | $bla_{OXA-71}$; *Mbl*; *Zn-dependent hydrolase*; $bla_{ADC-76}$ |
| **C-11** | ***Acinetobacter baumannii*** | $bla_{OXA-117}$; *Mbl*; *Zn-dependent hydrolase*; $bla_{ADC-238}$ |
| **C-12** | ***Acinetobacter baumannii*** | $bla_{OXA-68}$; *Mbl*; *Zn-dependent hydrolase*; $bla_{ADC-277}$; $bla_{ADC-76}$ |
| **C-14** | ***Acinetobacter baumannii*** | $bla_{OXA-117}$; Mbl; *Zn-dependent hydrolase*; $bla_{ADC-238}$ |
| **C-16** | ***Acinetobacter baumannii*** | $bla_{OXA-117}$; Mbl; *Zn-dependent hydrolase*; $bla_{ADC-238}$ |
| **C-18** | ***Acinetobacter baumannii*** | $bla_{OXA-117}$; Mbl; *Zn-dependent hydrolase*; $bla_{ADC-238}$ |
| **C-20** | ***Acinetobacter baumannii*** | $bla_{OXA-117}$; Mbl; *Zn-dependent hydrolase*; $bla_{ADC-238}$ |
| **C-21** | ***Acinetobacter baumannii*** | $bla_{OXA-117}$; *Mbl*; *Zn-dependent hydrolase*; $bla_{ADC-238}$ |

| | | |
|---|---|---|
| **C-22** | ***Acinetobacter baumannii*** | *bla*$_{\text{OXA-51}}$; *bla*$_{\text{A2}}$; *Zn-dependent_hydrolase*; *bla*$_{\text{ADC-165}}$ |
| **C-24** | ***Acinetobacter baumannii*** | *bla*$_{\text{OXA-71}}$; *Mbl*; *Zn-dependent hydrolase*; *bla*$_{\text{ADC-76}}$ |
| **C-25** | ***Acinetobacter baumannii*** | *bla*$_{\text{OXA-217}}$[1]; *Mbl*; *Zn-dependent hydrolase*; *bla*$_{\text{ADC-154}}$ |
| **C-11** | ***Acinetobacter nosocomialis*** | *bla*$_{\text{ADC-239}}$ |
| **C-12** | ***Acinetobacter nosocomialis*** | *Mbl*; *bla*$_{\text{ADC-130}}$ |
| **C-15** | ***Acinetobacter nosocomialis*** | *bla*$_{\text{OXA-23}}$; *bla*$_{\text{OXA-66}}$; *Mbl*; *Zn-dependent hydrolase*; *bla*$_{\text{ADC-30}}$ |
| **C-8** | *Chryseobacterium arthrosphaerae* | *bla*$_{\text{IND-7}}$ |
| **C-14** | *Chryseobacterium indologenes* | *bla*$_{\text{CIA-4}}$; *bla*$_{\text{IND-2b}}$ |
| **C-2** | *Comamonas testosteroni* | *bla*$_{\text{CARB-2}}$ |
| **C-14** | *Comamonas testosteroni* | *bla*$_{\text{CARB-2}}$ |
| **C-16** | *Elizabethkingia miricola* | *bla*$_{\text{B-38}}$; *bla*$_{\text{GOB-23}}$ |
| **C-21** | *Elizabethkingia miricola* | *bla*$_{\text{B-38}}$; *bla*$_{\text{GOB-23}}$ |
| **C-3** | ***Enterobacter hormaechei*** | *bla*$_{\text{ACT-24}}$; *bla*$_{\text{SHV-12}}$; *bla*$_{\text{TEM-1}}$ |
| **C-4** | ***Enterobacter hormaechei*** | *bla*$_{\text{ACT-24}}$ |
| **C-19** | ***Enterobacter hormaechei*** | *bla*$_{\text{ACT-24}}$; *bla*$_{\text{SHV-154}}$; *bla*$_{\text{TEM-1}}$ |
| **C-9** | ***Enterobacter roggenkampii*** | *bla*$_{\text{MIR-10}}$ |
| **C-16** | ***Enterobacter roggenkampii*** | *bla*$_{\text{MIR-5}}$ |
| **C-20** | ***Enterobacter roggenkampii*** | *bla*$_{\text{MIR-5}}$ |
| **C-21** | ***Enterobacter roggenkampii*** | *bla*$_{\text{MIR-5}}$ |
| **C-22** | ***Enterobacter roggenkampii*** | *bla*$_{\text{MIR-5}}$ |
| **C-8** | ***Escherichia coli*** | *AmpC1 Ecoli*; *Penicillin Binding Protein Ecoli*; *ampH Ecoli*; *bla*$_{\text{CTX-M-55}}$; *bla*$_{\text{EC-18}}$ |
| **C-9** | ***Escherichia coli*** | *AmpC1 Ecoli*; *Penicillin Binding Protein Ecoli*; *ampH Ecoli*; *bla*$_{\text{EC-13}}$ |
| **C-10** | ***Escherichia coli*** | *AmpC1 Ecoli*; *Penicillin Binding Protein Ecoli*; *ampH Ecoli*; *bla*$_{\text{CMY-2}}$; *bla*$_{\text{EC-18}}$ |
| **C-6** | ***Klebsiella pneumoniae*** | *ampH*; *bla*$_{\text{SHV-26}}$ |
| **C-9** | ***Klebsiella pneumoniae*** | *ampH*; *bla*$_{\text{SHV-36}}$ |
| **C-19** | ***Klebsiella pneumoniae*** | *ampH*; *bla*$_{\text{SHV-26}}$ |
| **C-20** | ***Klebsiella pneumoniae*** | *ampH*; *bla*$_{\text{SHV-26}}$ |
| **C-21** | ***Klebsiella pneumoniae*** | *ampH*; *bla*$_{\text{SHV-26}}$ |
| **C-8** | *Pandoraea apista* | *bla*$_{\text{OXA-153}}$ |
| **C-19** | *Pandoraea apista* | *bla*$_{\text{OXA-153}}$ |
| **C-20** | *Pandoraea apista* | *bla*$_{\text{OXA-153}}$ |
| **C-21** | *Pandoraea apista* | *bla*$_{\text{OXA-153}}$ |
| **C-6** | ***Serratia marcescens*** | *bla*$_{\text{SRT-3}}$ |

1. Low coverage hit

Notable Agar Findings

CHROMagar mSuperCARBA (mSC): The stated intended use of mSC agar is for qualitative detection of carbapenem-resistant Enterobacteriaceae (CPE) colonization in healthcare settings. One of the limitations of this agar is that bacteria with multidrug resistance or decreased membrane permeability may also grow, so detection is not limited strictly to CR organisms. Sequencing data showed many isolates that were phenotypically carbapenem-resistant did not have carbapenemase genes. Our dataset did, however, show a wide variety of other types of AR genes, the combination of which could account for the phenotypic carbapenem resistance (data not shown). Another limitation highlighted in the instructions for use (IFU) states that "Rarely, some VRE may grow in small blue colonies." In practice with wastewater samples, identification of *Enterococcus* spp. frequently occurred, with identification in 23.3% (17 of 73) of samples. *Enterococcus* spp. can often be differentiated from CPE on mSC based on size and color, as they typically appear in much smaller, lighter blue colonies than CPE colonies, which also sometimes have a purple halo. Another notable difference between the IFU and observations in wastewater samples is that *Aeromonas* spp. were observed to grow in an array of colors ranging between blue, gray, and purple. This contrasts with the IFU, which states that other gram-negative CP bacteria (i.e., those that are not Enterobacteriaceae or *Pseudomonas* spp.) should be colorless or have natural pigmentation. *C. freundii*, which should appear blue, was observed to have variable coloration. Most occurrences were variations of blue and purple coloration; however, there was one sample where *C. freundii* grew in a cream-colored colony on mSC. *E. coli* was only observed on mSC in one sample, and appeared cream-colored, in contrast to the expected dark pink to reddish color. Sequencing revealed that this isolate had several beta-lactam and efflux genes, but none of the "Big 5" carbapenemase genes (Supplemental Table 2; data not shown for efflux genes).

CHROMagar Acinetobacter + MDR Supplement (AC): The stated intended use of AC agar is for qualitative detection of *Acinetobacter* spp. in healthcare settings and clinical environments. The main challenge with using this agar for screening wastewater samples is there will be a variety of additional non-fermenting bacteria present in the samples, which can display similar coloration to *Acinetobacter* on AC agar. Collectively, 21 (32.9%) of these 73 samples showed a non-fermenter other than *Acinetobacter* spp. based upon the phenotypic characteristics using AC agar (of note, three samples had more than one non-fermenter on the same plate). Examples from this study are *Burkholderia* spp. (2.7%; 2 of 73 samples), *Chryseobacterium* spp. (6.9%; 5 of 73 samples), *Comamonas* spp. (6.9%; 5 of 73 samples), *Elizabethkingia* spp. (4.1%; 3 of 73 samples), *Pandoraea* spp. (9.6%; 7 of 73 samples), and *Sphingobacterium* spp. (2.7%; 2 of 73 samples). These genera showed a range of coloration; some appeared clear or clear with a red bullseye, while others grew various shades of red. Distinction between *Acinetobacter* spp. and other bacteria that also display red coloration on this agar becomes easier with experience; however, confirmatory tests should be used for identification.

CHROMagar Pseudomonas (PC): To efficiently identify *Pseudomonas* spp. in wastewater samples, PC was tested, as the translucent/cream color of *Pseudomonas* spp. typically observed on mSC makes it difficult to isolate from amongst the variety of other bacteria that may grow in similar coloration. Two problems were observed when using this agar with wastewater samples. Firstly, due to the absence of antibiotics in this agar, much less inhibition was observed compared to the other agars used, which resulted in overgrowth, even at higher dilutions. Secondly, *Pseudomonas* spp. was identified in three of 36 (8.3%) blue colonies from PC (only *Pseudomonas* spp. should appear as blue-green colonies per the IFU). The other genera identified with blue coloration on PC agar were *Aeromonas* spp. (19.4%, 7 of 36), *Enterobacter* spp. (41.7%, 15 of 36), *Escherichia* spp. (2.8%, 1 of 36), *Klebsiella* spp. (13.9%, 5 of 36), *Kluyvera* spp. (2.8%, 1 of 36),

*Shimwellia* spp. (8.3%, 3 of 36), and isolates that were not identifiable by the MALDI database (2.8%, 1 of 36).

PC agar is not selective enough for use with wastewater, as the vast microbial community overgrows in the absence of antimicrobial pressure, outcompeting *Pseudomonas* spp., even at higher dilutions. The presence of non-*Pseudomonas* spp. observed in various shades of blue made identifying true *Pseudomonas* spp. difficult.
